# Serum Total Immunoglobulin-E Levels and Early Loss of eGFR in Individuals at Risk of Mesoamerican Nephropathy: A Nested Case-Control Analysis from a Population Representative Follow-Up Study

**DOI:** 10.64898/2026.02.27.25342157

**Authors:** Ben Caplin, Sanskriti Agarwal, Alexandra Day, Ali Al-Rashed, Amin Oomatia, Marvin Gonzalez-Quiroz, Neil Pearce

## Abstract

**Introduction:** There remains considerable debate as to the cause of the epidemic of Mesoamerican Nephropathy (MeN). We have previously reported early loss of estimated glomerular filtration rate (eGFR) as a surrogate for disease onset in a population-representative cohort study of young-adults at risk of disease from Northwest Nicaragua. Using a nested case-control approach we analysed urine and serum proteins surrounding this timepoint with the aim of gaining insight into the primary disease aetiology.

**Methods:** We conducted label-free ultra high-performance liquid chromatography mass-spectrometry based proteomics using urine samples collected at the study visit before, and at, first observed eGFR loss amongst cases and compared results to matched controls. We then performed direct protein measurements in a discovery cohort followed by quantification of serum total immunoglobulin E (_st_IgE) at multiple timepoints in a replication cohort.

**Results:** Proteomic analysis demonstrated no differences in the levels of any single protein between cases and controls (n=25 each), at either timepoint, after correction for multiple comparisons. However, functional enrichment analysis demonstrated upregulation of adaptive immune pathways amongst cases. Direct measurements in the discovery cohort using high-sensitivity PCR-based immunoassay (n=21 controls, 19 cases) demonstrated higher _st_IgE in cases at the study visit immediately prior to first observed eGFR loss (mean difference 810kU/L, 95% confidence interval (CI): 162-1457kU/L). In the replication cohort (n=22 cases, 21 controls) an _st_IgE level >500kU/L measured by electrochemiluminescence in study samples from any timepoint in the 3 years prior to the first observed loss of eGFR was independently associated with case status when compared to samples from controls at matched visits (adjusted Odds Ratio: 8.1, 95% CI: 1.4-47.8).

**Discussion:** A high level of _st_IgE precedes loss of eGFR in those at risk of MeN. Understanding what leads to this rise is likely to be key to understanding the cause of the MeN epidemic.

**Lay Summary:** Mesoamerican nephropathy describes an epidemic-level chronic kidney disease impacting rural working age adults in Central America. Although a number of exposures, including occupational heat exposure, have been proposed the cause of the epidemic, there remains much debate as to the primary aetiology of the disease. In this study we interrogated urine and blood samples from individuals from affected communities at risk of disease both before and after they develop kidney dysfunction. Using two different approaches, analysis of both urine and blood samples provide evidence of upregulation of immunoglobulin-E (IgE) related pathways in the 2-3 years before individuals develop evidence of kidney disease. Infections (particularly those involving parasites) and allergic reactions, but not heat exposure, have been reported to increase IgE levels. Going forwards, understanding the cause of this increase in IgE in individuals at risk of disease is likely to provide insight into the cause of Mesoamerican Nephropathy epidemics.

## Introduction

Mesoamerican Nephropathy (MeN) is the term used to describe the epidemic-level chronic kidney disease of unknown cause impacting several regions in Central America[1]. MeN primarily affects rural, working age adults and is reported as a leading cause of years of life lost amongst this group in El-Salvador and Nicaragua[2]. Despite two decades of research and the identification of a number of risk factors, the primary cause of epidemic-level disease in this region is uncertain.

One reason for this is that much of the analytical epidemiology has interrogated risk factors for established disease (i.e., chronic kidney disease, CKD, or an estimated glomerular filtration rate, eGFR<60mL/min/1.73m^2^), whereas this outcome, in common with other causes of CKD, will represent a composite of initiating causes and exacerbating factors. Given the burden of generic CKD exacerbating factors (e.g., nephrotoxic drug exposure or episodes of heat exposure and dehydration) in settings impacted by MeN these analytical approaches may not capture associations with exposures linked to the initiation of disease (i.e. prior to the decline in eGFR).

Over the last decade, we have been conducting a population-representative follow-up study in initially apparently healthy young adults living in the at risk communities in Northwest Nicaragua[3]. Serial eGFR testing allows us to observe early loss of kidney function within individuals whilst the eGFR is in the normal or near-normal range. We have previously reported the use of this early eGFR loss as a surrogate for disease onset[4]. To better understand the primary cause of disease, we conducted a nested case-control study initially characterising the urinary proteome around this time point. We then conducted follow-up testing of blood and urine before confirming the key findings in a replication sample with the aim of providing clues as to disease aetiology.

## Short Methods

Using a nested case-control approach, we used serum and urine from incident cases experiencing early loss of eGFR, and controls, with stable eGFR, matched for age and sex, identified from our follow-up study (total n=771)[4]. Initially we conducted label free data independent acquisition-based ultra high-performance liquid chromatography mass-spectrometry proteomics[5]. This analysis used urine from cases, both from the visit before and the visit of first observed eGFR loss, with control samples from matched timepoints. Following this we assessed seropositivity to nephrotropic organisms. We then divided the cohort into two and performed direct immunoglobulin and chemokine measurements in serum and urine in cases and controls from a discovery sample, alongside participants classified as established cases. Finally, in a replication sample, we quantified serum total immunoglobulin E (_st_IgE) in blood samples collected at multiple time points using external quality-assured electrochemiluminescence. Full methods are described in the Supplementary Appendix.

## Results

Urinary proteomics was conducted in a total of 96 urine samples (Supplementary Table 1). Following protein identification, imputation of missing values and correction for multiple comparisons, no difference in any individual protein was identified between cases and controls, at either the visit before (Figure 1A) or the visit of (Supplementary Figure 1) first observed eGFR loss in cases. Functional enrichment analysis using the Reactome canonical pathways database[6] demonstrated overrepresentation of adaptive immune pathways in cases at the visit immediately prior to first observed eGFR loss (Figure 1B). Comparisons at the visit of first observed eGFR loss (Supplementary Table 2) and analyses using other databases (i.e., GO Molecular Function Ontology, Supplementary Table 3) showed similar results. No differences between cases and controls were seen in ELISA reactivity for *Leptospira sp*. (Supplementary Figure 2) and hantavirus seropositivity was rare (Supplementary Table 4).

**Figure:**
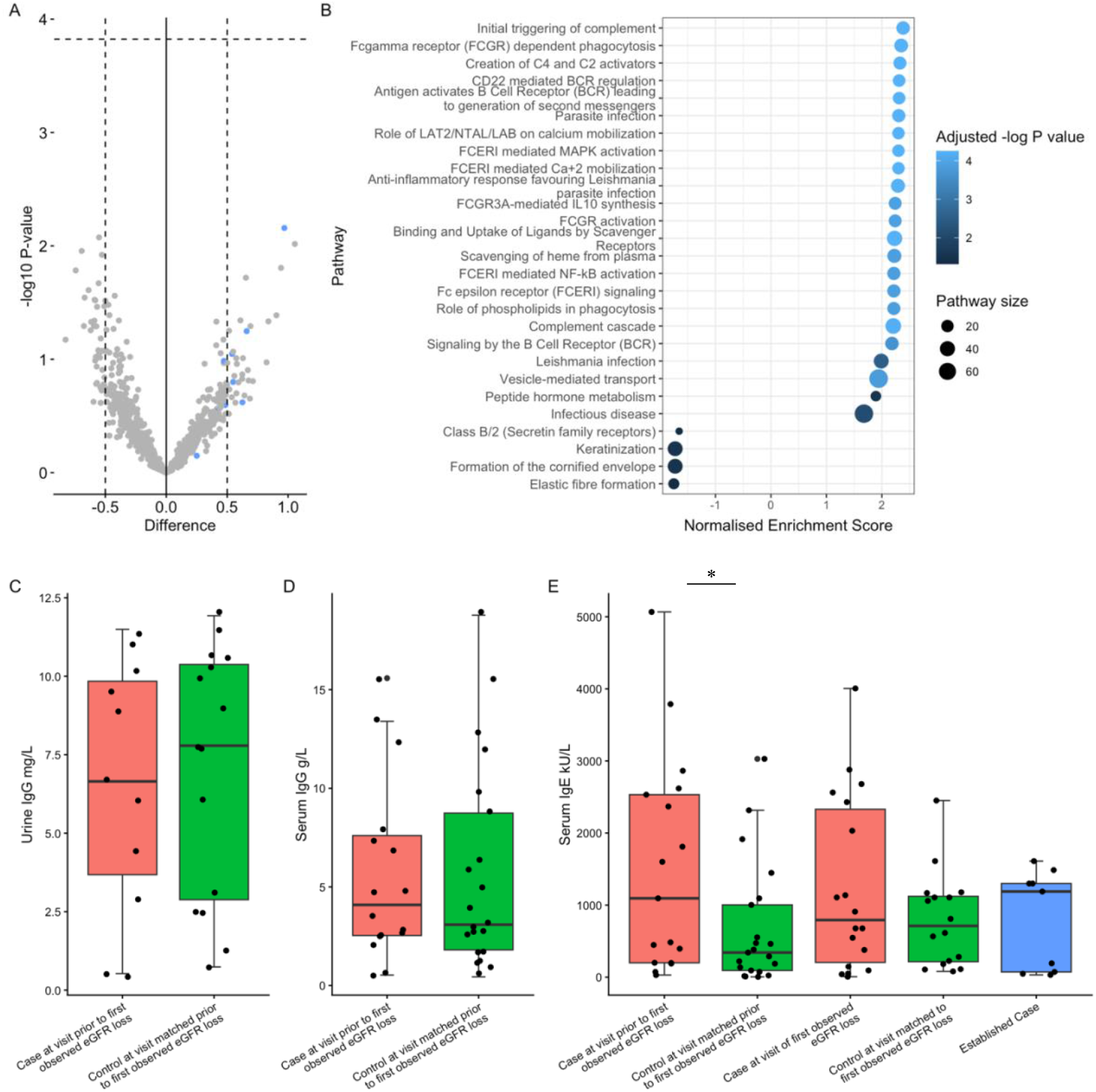
Proteomics analysis and immunoglobulin measurements in the discovery cohort. **(A)** Volcano plot of differences in protein abundances between cases versus controls at the time point immediately prior to first observed eGFR loss. Dashed horizontal line reflects threshold for significance after adjustment for multiple comparisons using the Benjamini-Hochberg procedure. **(B)** Pathway enrichment analysis in cases versus controls at the time point immediately prior to early loss of eGFR using gene sets listed in the reactome database. Only pathways with a Benjamini-Hochberg adjusted p-value <0.05 are shown. **(C)** Urine, and **(D)** serum, immunoglobulin G levels quantified by enzyme-linked immune-assay in cases (red) at the time point immediately prior to first observed eGFR loss and controls (green) matched for study visit. No differences between groups were identified. **(E)** Serum total IgE levels measured by high-sensitivity PCR based method in cases (red) at time points immediately prior to and at time of first observed eGFR loss and controls (green) matched for study visit, as well as the established case group (blue). \**P*=0.012 for a difference between incident cases and controls at time point immediately prior to first observed loss of eGFR using a mixed model including case status and time point. All other comparisons non-significant.

Characteristics of the matched nested case-control discovery sample used for direct immunoglobulin and chemokine measures, as well as the group with established disease, are described in Table 1. No differences in serum or urine total IgG were observed between cases and controls at the timepoint immediately prior to first observed eGFR loss in cases (Figure 1C and 1D). We were unable to detect urinary IgE using high-sensitivity PCR based methods but overall _st_IgE levels were high, and cases had a higher _st_IgE at the time point prior to first observed eGFR loss versus controls sampled at the same timepoint (mean difference 810kU/L, 95% confidence interval (CI): 162-1457kU/L; Figure 1E). Urinary IL-4 was undetectable using high sensitivity enzyme-linked immunoassay, but we did observe borderline evidence of an increase in urinary Chemokine (C-X-C motif) ligand-9 in cases versus controls across both time points (Supplementary Figure 3).

**Table 1.** Description of the study samples for IgE analysis.

|  | Discovery Sample |  |  | Replication Sample |  |
| --- | --- | --- | --- | --- | --- |
|  | Control | Incident Case | Established Case | Control | Incident Case |
| N | 21 | 19 | 9 | 21 | 22 |
| Sex |  |  |  |  |  |
| Male | 21 (100.0%) | 19 (100.0%) | 9 (100.0%) | 14 (66.7%) | 15 (68.2%) |
| Female |  |  |  | 7 (33.3%) | 7 (31.8%) |
| Age | 24.9 (4.5) | 24.0 (3.4) | 24.7 (3.5) | 25.3 (4.6) | 26.5 (3.6) |
| Sugarcane Work Ever |  |  |  |  |  |
| No | 4 (19.0%) | 1 (5.3%) | 2 (22.2%) | 13 (61.9%) | 9 (40.9%) |
| Yes | 17 (81.0%) | 18 (94.7%) | 7 (77.8%) | 8 (38.1%) | 13 (59.1%) |
| Community |  |  |  |  |  |
| 1 | 2 (9.5%) | 0 (0.0%) | 0 (0.0%) | 2 (9.5%) | 3 (13.6%) |
| 2 | 3 (14.3%) | 2 (10.5%) | 0 (0.0%) | 2 (9.5%) | 1 (4.5%) |
| 3 | 3 (14.3%) | 3 (15.8%) | 0 (0.0%) | 1 (4.7%) | 1 (4.5%) |
| 4 | 2 (9.5%) | 2 (10.5%) | 1 (11.1%) | 1 (4.7%) | 1 (4.5%) |
| 5 | 2 (9.5%) | 2 (10.5%) | 1 (11.1%) | 0 (0.0%) | 0 (0.0%) |
| 6 | 7 (33.3%) | 7 (36.8%) | 0 (0.0%) | 1 (4.7%) | 1 (4.5%) |
| 7 | 2 (9.5%) | 3 (15.8%) | 3 (33.3%) | 2 (9.5%) | 1 (4.5%) |
| 8 | 0 (0.0%) | 0 (0.0%) | 2 (22.2%) | 4 (19.0%) | 4 (18.2%) |
| 9 | 0 (0.0%) | 0 (0.0%) | 2 (22.2%) | 5 (23.8%) | 5 (22.7%) |
| 10 | 0 (0.0%) | 0 (0.0%) | 0 (0.0%) | 1 (4.7%) | 1 (4.5%) |
| eGFR mL/min/1.73m<sup>2</sup>, mean (SD) |  |  |  |  |  |
| Baseline | 122.6 (9.0) | 116.8 (12.1) |  | 118.3 (12.2) | 117.2 (9.7) |
| Visit prior to eGFR loss | 118.4 (8.2)* | 112.9 (8.4) |  | 117.9 (6.7)* | 111.1 (6.2) |
| Visit following eGFR loss | 115.8(8.9)* | 84.0 (6.7) | 55.5 (9.2)<sup>†</sup> | 112.2 (10.4)* | 85.1 (5.4) |
| Interval from baseline to eGFR loss, years, median (range) |  | 1.5 (1 to 5) |  |  | 3 (1 to 5) |
| Number of IgE measures per individual, median (range) | 2 (1 to 2) | 2 (1 to 2) | 1 (1) | 5 (2 to 5) | 4 (2 to 5) |

\*For controls (with stable eGFR), eGFR values are from the matched study visits defined by the eGFR in cases.
<sup>†</sup>eGFR in established disease group is at time of sampling.

We then aimed to confirm our findings in a replication sample. We measured _st_IgE at multiple time points using electrochemiluminescence, in serum samples collected up to 36-months prior to, and 12 months following, first observed eGFR loss in cases and compared to age- and sex- and study visit-matched controls (Table 1). Across all time points taken together, cases had a higher _st_IgE, with mean estimates from the multilevel model of 631.4kU/L (95%CI: 458.2 to 804.5kU/L) versus 262.0kU/L (95%CI: 85.9 to 438.0kU/L) in controls. Trajectories of _st_IgE in cases varied over time, with persistently high levels in some individuals and levels peaking at times prior to first observed eGFR loss in others (Supplementary Figure 4). Overall, an _st_IgE level >500kU/L at, or any time in the 3 years prior to, first observed loss of eGFR (which occurred in 14 cases and 5 controls) was strongly and independently associated with case status when compared to matched controls during the same period after adjustment for demographic and occupational variables (Table 2). Finally, to ensure our findings weren’t the consequence of the statistical methods used to define case status, we examined direct longitudinal associations between _st_IgE and eGFR in the total replication sample. This demonstrated negative associations between stIgE and eGFR up to 2 visits following _st_IgE measurement (Supplementary Figure 5).

**Table 2.**
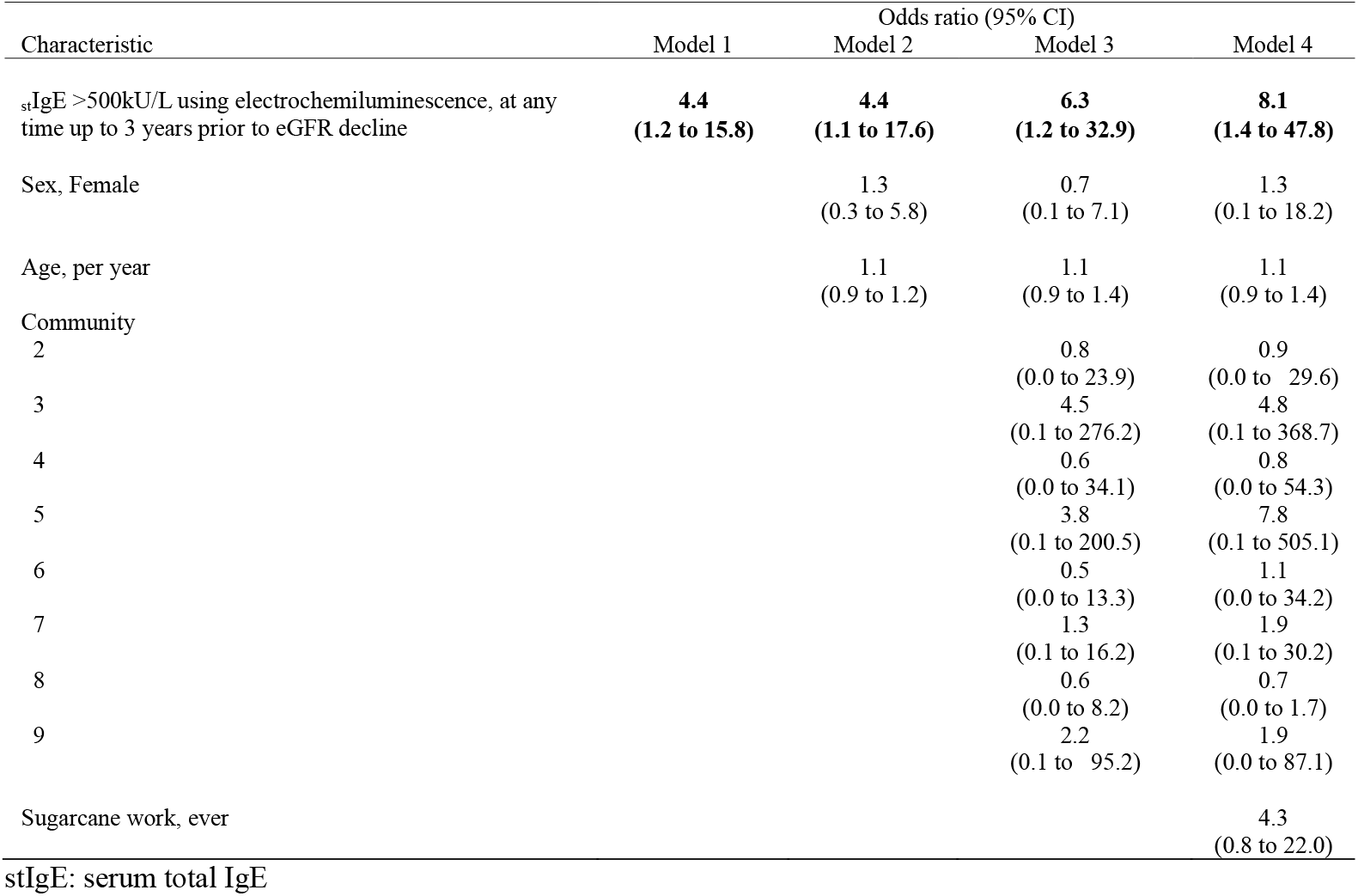
Association between high serum total IgE and case status in the replication sample adjusted for potential confounders.

## Discussion

We interrogated the urinary proteome and conducted confirmatory analyses in individuals from a population at risk of MeN. We identified an adaptive immune response, but in common with previous studies without evidence of involvement of classic nephrotropic infections[7]. However, we observe a markedly elevated _st_IgE in individuals who go on to experience decline in eGFR. The population-representative cohort, orthogonal methods (unbiased urinary proteomics and direct immunoassays), replication sample, absence of evidence of confounding by demographic or occupational factors, and time course of the associations (i.e., at, or up to 3 years prior to, first observed eGFR loss) support an important relationship between the elevated _st_IgE and the cause of early kidney dysfunction in this population. In turn, this strongly suggests this IgE response is related to the primary aetiology of MeN.

Despite the above strengths these studies have limitations. Firstly, our case definitions, although likely capturing early disease more effectively than alternative approaches, are nonetheless likely to be subject to misclassification as the presence of substantial renal reserve in the young adults included in the study, means significant kidney injury may be present in the absence of any change in eGFR[8]. In turn this means that using eGFR measures to examine temporal associations with exposures related to disease onset remains prone to error. Secondly, we have only measured total IgE. In common with many poor rural populations in tropical regions, _st_IgE levels were high in the overall study population, likely reflecting exposure to geohelminths[9]. Hence the _st_IgE levels we have observed associated with disease onset represent an increase above an already high baseline. Our studies do not provide insight into the exposure that leads to this increase, and outside of the atopy field there are no routinely available standardised methods to characterise the specificity of IgE responses[10]. Unfortunately, we have been unable to measure eosinophil levels given the pre-analytical challenges associated with differential blood counts. Finally, we used two different methods to quantify _st_IgE, meaning the results from the discovery and replication cohorts, although consistent, cannot be combined.

Common causes of high IgE levels include: (i) infection, classically metazoan parasites, although increases in response to other microorganisms (e.g., viruses and fungi) are also well described; (ii) atopy, where IgE reactivity occurs to common environmental antigens; and (iii) malignant, genetic and autoimmune disorders. Individuals in MeN affected populations do not describe typical symptoms of allergic disease and atopy is not classically associated with kidney disease. Raised levels of IgE are described in cases of (specifically drug-induced) acute interstitial nephritis (AIN)[11, 12], and IgE deposition has been observed in biopsies with other forms of tubulointerstitial inflammation[13] but any mechanistic role for IgE in mediating injury remains uncertain. An alternative explanation for our findings is that the IgE response is a surrogate for exposure to a cause of kidney injury which acts through a cell-mediated, or possibly toxin induced, mechanism. For example, there are descriptions of classical (presumed cell-mediated) AIN with parasitic infections[14]. Relatedly, the relevance of our findings to the series of patients with AIN reported from hospitals in the region[15] is unclear. It is possible that the elevated _st_IgE is a more general marker of autoimmunity or causes renal injury independent of any specific exposure. This seems unlikely, however, given the magnitude of the association and absence of evidence of more generalised autoimmunity. Furthermore, there are no reports of a substantial burden of kidney disease in other groups with high circulating levels of total IgE.

In conclusion we have identified elevated _st_IgE preceding loss of eGFR over the following 2-3 years in a population at risk of MeN with normal baseline kidney function. Understanding the exposure associated with this rise in IgE seems critical to gaining insight into the cause of the MeN epidemic.

## Data Availability

Data described in this manuscript will be made available on reasonable request to the authors.

## Data Sharing Statement

Proteomics data are available on ProteomeXchange via the PRIDE partner repository: https://www.ebi.ac.uk/pride (identifier PXD070158). Other data described in this manuscript will be made available on reasonable request to the authors.

## Author Contributions

BC and NP conceived and designed the cohort study along with MGQ, who led the field team in Nicaragua. BC conceived and planned this sub-study, analysed the proteomic data and wrote the first draft of the manuscript. SA, AO, AD and AA-R analysed the samples and results. All authors read and approved the final version of the manuscript.

## Acknowledgements

The authors thank the study participants and leaders from the study communities as well as the Nicaraguan fieldwork team, without whom, none of this work would be possible. We thank Steven Lynham and the team at the Kings College London Centre of Excellence for Mass Spectrometry. The authors also wish to thank Eva Smpokou and Donna Davoren for technical and administrative assistance. The authors also acknowledge the contributions of FNE International for practical support, members of La Isla Network for their input at the initial stages of this work, and the Getting to the Cause of CKDu Study steering group.

