## Supplemental Material for "Serum Total Immunoglobulin-E Levels and Early Loss of eGFR in Individuals at Risk of Mesoamerican Nephropathy: A Nested Case-Control Analysis from a Population Representative Follow-Up Study"

**Supplementary Material**

**Supplementary Methods**

*Cohort and samples*

Approval for this work was received from the institutional review boards at Universidad Nacional Autonoma de Nicaragua, Leon (Acta No.116, Ano 2014; Acta No.71, Ano 2018), the London School of Hygiene and Tropical Medicine (Ref: 8643) and University College London (Ref: 14175). All cohort participants provided written informed consent.

The study cohort and methods by which incident cases and matched controls were selected have been previously described[1, 2]. Briefly, the study cohort was recruited from 11 communities in Northwest Nicaragua from 2014 onwards. Individuals, aged 18-30, were invited to self-report demographics, medical history, occupation and occupational exposures and symptoms as well as undergo clinical measurements and blood and urine sampling. Study visits initially occurred 6-monthly for two years and annually thereafter. Incident cases were defined as individuals who were observed to have a Markov probability of >0.5 of moving from a healthy to an unhealthy eGFR distribution between two study visits. The latter of these two study visits was defined as the time point of early eGFR loss. Control samples with stable eGFR were matched for study visit. Not all samples were available at all time points for all analyses. Replacement samples were not used leading to unbalanced groups at each time point.

Serum (separated in the field) and urine were initially stored on ice during field collection and then transferred to -20°C storage in Nicaragua for up to 4 weeks before being transferred to the UK on dry ice. Once in the UK all samples were stored at -80°C until analysis.

*Proteomics*

We conducted label-free proteomics using a data-independent acquisition approach. Protease inhibitors (Roche cOmplete ULTRA; cat no: 05892970001) were added to all samples on thawing. Samples were normalised to 100ug total protein, cysteine residues were reduced and derivatised to form stable carbamidomethyl derivatives before trypsin digestion overnight at 37C. C18 spin column clean-up was performed prior to resuspension (0.5mcg/mcL) and 2mcL analysed by LC-MS/MS.

Chromatographic separation was performed using a U3000 UHPLC NanoLC system with peptides were resolved by reversed phase chromatography on a 75mm C18 Pepmap column using a stepwise linear gradient of 0.1% formic acid in water and 80% acetonitrile in 0.1% formic acid. The gradient was delivered to elute the peptides at a flow rate of 250nl/min over 60 min.

The eluate was ionised with an Orbitrap Eclipse mass spectrometer (ThermoFisher Scientific) operating Xcalibur v4.3.69.37 using a data-independent acquisition (DIA) method, higher-energy collision dissociation with a collision energy of 30% for MS/MS fragmentation. The full MS scan was obtained using a precursor mass range of m/z 400-900 with orbitrap resolution of 60,000, AGC 300% (1.200e6) and a maximum injection time: Auto. DIA was used for the MS2 scan, with an m/z isolation window of 12, 1 overlap, and 42 scan events with scan range m/z 145-1450. The MS2 scan used an AGC setting of 800% with absolute AGC value: 4.000e5, and maximum injection time: auto with loop control: All.

We used a data independent acquisition method with the DIA-NN pipeline[3]. Raw mass spectrometry data were processed against the Uniprot Human Taxonomy database (uniprotkb_human_reviewed_2024_03_09) with a 1% FDR. Precursor ion generation was performed using a library-free search based on deep learning-based spectra, retention times and IMs prediction option being switched on and calculated against the database fasta file. Trypsin/P was selected with 2 missed cleavages and Ox (M) and carbamidomethylation (C) included. Peptide length range (6-30), peptide charge (1-4), precursor m/z range (300-1800) and fragment ion m/z range (145-1800) also selected for searching. Cross-run normalisation was based on RT-dependent with match between runs (MBR) switched on. The mass spectrometry proteomics data have been deposited to the ProteomeXchange Consortium via the PRIDE[4] partner repository with the dataset identifier PXD070158.

Extracted protein level relative abundances were exported to Rstudio and a probabilistic dropout approach used to identify differentially abundant proteins using the R package ProDA[5]. Comparisons were made between case and matched control samples separately at the two time points. P-values were adjusted for multiple comparisons using the Benjamini-Hochberg procedure with an adjusted p-value <0.05 accepted as significant.

Pathway enrichment analysis was then performed using both the Reactome[6] and Gene ontology: Molecular function[7] databases using the R package fgsea[8]with default settings and Benjamini-Hochberg adjusted p-values and an adjusted p-value <0.05 accepted as significant.

*Infectious serology*

Anti-Leptospira IgG and anti-hantavirus IgG were measured using Abcam Leptospira IgG ELISA kit (cat no: ab247199) and Euroimmun anti-hantavirus IgG 'Americas' ELISA (cat no: EI03833; reported reactivity to both *Andes* and *Sin Nombre* strains), respectively according to manufacturer's instructions.

*Immunoglobulin and chemokine measures*

IgG was measured using Abcam human IgG ELISA kit (cat no: ab195215); IgE by ThermoFischer Scientific Human IgE ProQuantum Immunoassay Kit (cat no: A44690); Il-4 by Invitrogen High Sensitivity Human IL-4 ELISA Kit (BMS225-2HS); and CXCL9 by Abcam Human CXCL9 ELISA kit (cat no: ab219047).

In the replication sample, $_{st}$IgE was quantified using an externally quality assured electrochemiluminescence assay (Roche Elecsys IgE II).

*Other statistical methods*

Descriptive statistics for the discovery and replication samples were presented using frequencies (and percentages), means (and standard deviations) or medians (and either range or interquartile range) as most appropriate. Tests of between group difference in IgG at the visit prior to eGFR decline were conducted with T-tests. For the analysis of anti-Leptospira IgG, CXCL9 and $_{st}$IgE levels across multiple time points we used a mixed model with a random intercept. Antibody (or CXCL9) measurements were nested within individuals with inclusion of both case status and (except in the replication sample $_{st}$IgE analysis where measures occurred at up to 5 time points) a case status*time point interaction

To understand the association between case status and raised $_{st}$IgE within the 3 years prior to loss of eGFR we used logistic regression models. After examining the crude association, we adjusted the models for *a priori* predicted confounders: age, sex, community and occupation (in the form of sugarcane work). Lastly to examine the temporal relationships between $_{st}$IgE and eGFR we used the $_{st}$IgE coefficients from mixed models modelling eGFR as an outcome over time. We separately modelled eGFR at the visit prior to each $_{st}$IgE measurement, at the same time as each $_{st}$IgE measurement, and for each of the three visits following each $_{st}$IgE measurement. These models used $_{st}$IgE measurements nested within individuals and adjusted for age, sex, study visit and follow-up time and inclusion of a random intercept and slope for follow-up time.

All analysis was conducted in Stata version 19 (StataCorp, College Station, Texas) and Rstudio version 2025.09.1 (Posit Software PBC and R Foundation for Statistical Computing, Vienna, Austria)

**Supplementary Figure Legends**

**Supplementary Figure 1 - Volcano plot of differences in protein abundances between cases versus controls at the time of first observed eGFR loss**

Dashed horizontal line reflects threshold for significance after adjustment using the Benjamini-Hochberg procedure

**Supplementary Figure 2 – Results from Leptospira ELISA**

Controls (green) and cases (red) at visits before (-1) and immediately following (0) early loss of eGFR. No differences between cases and controls (n=57 each) at either visit. Dashed line: threshold for borderline positive result; Solid line threshold for confirmed positive result.

**Supplementary Figure 3 - Urinary CXCL9 levels in incident cases and controls in the discovery cohort.**

Controls (green) and cases (red) at visits before (-1) and immediately following (0) early loss of eGFR. $P$=0.055 for a difference between cases and controls across both time visits using linear mixed model.

**Supplementary Figure 4 - Spaghetti plots of serum total IgE trajectories in the replication cohort**

Serum total IgE by interval from timepoint of early eGFR loss in cases (with controls matched for time point). Controls (0) and cases (1).

**Supplementary Figure 5 - Association between serum total IgE and eGFR by different time lags**

Difference in eGFR by interval from IgE measurement in the overall replication cohort (cases and controls combined). Negative values on the x-axis represent eGFR values before the IgE measure and positive values eGFR measures following the IgE measurement. Coefficients and 95% confidence intervals derived from mixed models adjusted for age, sex, study visit and follow-up time.

**Supplementary Table 1 - Description of samples for proteomics**

| | Controls (n=25) | | Incident cases (n=25) | |
|---|---|---|---|---|
| | Visit prior to eGFR decline* | Visit following eGFR decline* | Visit prior to eGFR decline | Visit following eGFR decline |
| Sample n | 24 | 22 | 24 | 25 |
| Age, mean (SD) | 24.96 (4.60) | 25.68 (4.76) | 24.19 (3.52) | 25.08 (3.40) |
| Sex, Male (%) | 24 (100.0) | 22 (100.0) | 24 (100.0) | 25 (100.0) |
| eGFR mL/min/1.73m$^2$, mean (SD) | 117.58 (7.55) | 115.94 (9.01) | 111.31 (8.23) | 84.42 (7.69) |

*For controls (with stable eGFR), these are the matched study visits defined by the eGFR in the cases

**Supplementary Table 2 – Pathway enrichment analysis using Reactome at the visit at the time of first observed eGFR loss**

| pathway | pval | adjp | log2error | ES | NES | size | gsname |
|---|---|---|---|---|---|---|---|
| 2730905 | 4.78E-07 | 3.35E-05 | 0.659444 | 0.796155 | 2.462544 | 20 | REACTOME ROLE OF LAT2 NTAL LAB ON CALCIUM MOBILIZATION |
| 2871796 | 4.78E-07 | 3.35E-05 | 0.659444 | 0.796155 | 2.462544 | 20 | REACTOME FCERI MEDIATED MAPK ACTIVATION |
| 2871809 | 4.78E-07 | 3.35E-05 | 0.659444 | 0.796155 | 2.462544 | 20 | REACTOME FCERI MEDIATED CA 2 MOBILIZATION |
| 2454202 | 3.57E-07 | 3.35E-05 | 0.674963 | 0.752215 | 2.470226 | 25 | REACTOME FC EPSILON RECEPTOR FCERI SIGNALING |
| 2871837 | 3.57E-07 | 3.35E-05 | 0.674963 | 0.752215 | 2.470226 | 25 | REACTOME FCERI MEDIATED NF KB ACTIVATION |
| 983705 | 1.05E-06 | 4.58E-05 | 0.643552 | 0.720429 | 2.411969 | 27 | REACTOME SIGNALING BY THE B CELL RECEPTOR BCR |
| 5690714 | 9.35E-07 | 4.58E-05 | 0.659444 | 0.77661 | 2.440602 | 21 | REACTOME CD22 MEDIATED BCR REGULATION |
| 983695 | 9.35E-07 | 4.58E-05 | 0.659444 | 0.77661 | 2.440602 | 21 | REACTOME ANTIGEN ACTIVATES B CELL RECEPTOR BCR LEADING TO GENERATION OF SECOND MESSENGERS |
| 2029481 | 2.09E-05 | 7.30E-04 | 0.57561 | 0.697297 | 2.239731 | 23 | REACTOME FCGR ACTIVATION |
| 9664323 | 2.09E-05 | 7.30E-04 | 0.57561 | 0.697297 | 2.239731 | 23 | REACTOME FCGR3A MEDIATED IL10 SYNTHESIS |
| 9664407 | 2.52E-05 | 8.00E-04 | 0.57561 | 0.673162 | 2.210621 | 25 | REACTOME PARASITE INFECTION |
| 2029485 | 9.63E-05 | 2.81E-03 | 0.538434 | 0.652634 | 2.116462 | 24 | REACTOME ROLE OF PHOSPHOLIPIDS IN PHAGOCYTOSIS |
| 166786 | 1.12E-04 | 3.01E-03 | 0.538434 | 0.647287 | 2.099121 | 24 | REACTOME CREATION OF C4 AND C2 ACTIVATORS |
| 166658 | 1.85E-04 | 4.62E-03 | 0.518848 | 0.515459 | 1.945674 | 45 | REACTOME COMPLEMENT CASCADE |
| 166663 | 2.09E-04 | 4.88E-03 | 0.518848 | 0.617369 | 2.06693 | 27 | REACTOME INITIAL TRIGGERING OF COMPLEMENT |
| 2168880 | 2.98E-04 | 6.52E-03 | 0.498493 | 0.560168 | 1.942028 | 31 | REACTOME SCAVENGING OF HEME FROM PLASMA |
| 9006934 | 3.81E-04 | 7.84E-03 | 0.498493 | -0.63742 | -1.78228 | 34 | REACTOME SIGNALING BY RECEPTOR TYROSINE KINASES |
| 2029480 | 6.31E-04 | 1.23E-02 | 0.477271 | 0.564479 | 1.924252 | 29 | REACTOME FCGAMMA RECEPTOR FCGR DEPENDENT PHAGOCYTOSIS |
| 6803157 | 1.01E-03 | 1.87E-02 | 0.45506 | 0.60669 | 1.876521 | 20 | REACTOME ANTIMICROBIAL PEPTIDES |
| 198933 | 1.70E-03 | 2.84E-02 | 0.45506 | 0.440867 | 1.700071 | 49 | REACTOME IMMUNOREGULATORY INTERACTIONS BETWEEN A LYMPHOID AND A NON LYMPHO ID CELL |
| 2173782 | 1.71E-03 | 2.84E-02 | 0.45506 | 0.469545 | 1.74907 | 42 | REACTOME BINDING AND UPTAKE OF LIGANDS BY SCAVENGER RECEPTORS |
| 9662851 | 2.20E-03 | 3.50E-02 | 0.431708 | 0.502174 | 1.740972 | 31 | REACTOME ANTI INFLAMMATORY RESPONSE FAVOURING LEISHMANIA PARASITE INFECTION |

**Supplementary Table 3 – Pathway enrichment analysis using Gene Ontology Molecular Function at the visit before early loss of eGFR**

| pathway | pval | adjp | log2error | ES | NES | size | gsname |
|---|---|---|---|---|---|---|---|
| GO:0003823 | 6.39E-05 | 0.015785 | 0.538434 | 0.581865 | 2.045041 | 42 | GOMF ANTIGEN BINDING |
| GO:0060089 | 1.18E-04 | 0.015785 | 0.538434 | -0.49828 | -1.74749 | 100 | GOMF MOLECULAR TRANSDUCER ACTIVITY |

**Supplementary Table 4 - Hantavirus ELISA results**

| | Controls (n=40) | | Incident cases (n=39) | |
|---|---|---|---|---|
| | Visit prior to eGFR decline* | Visit following eGFR decline* | Visit prior to eGFR decline | Visit following eGFR decline |
| Negative | 39 | 23 | 38 | 23 |
| Borderline | 1 | 1 | 0 | 0 |
| Positive | 0 | 0 | 1 | 0 |

**Supplementary Figure 1 - Volcano plot of differences in protein abundances between cases versus controls at at the time of first observed eGFR loss**

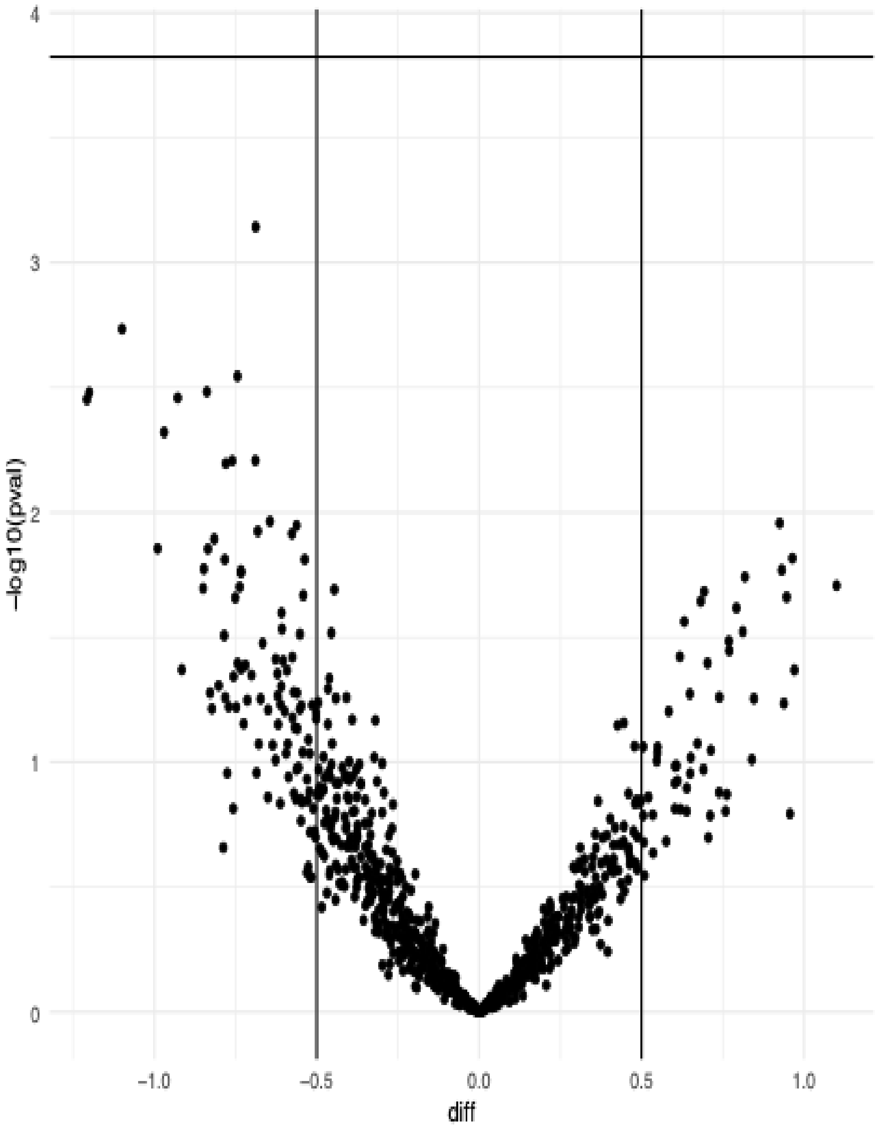

**Supplementary Figure 2 – Results from Leptospira ELISA**

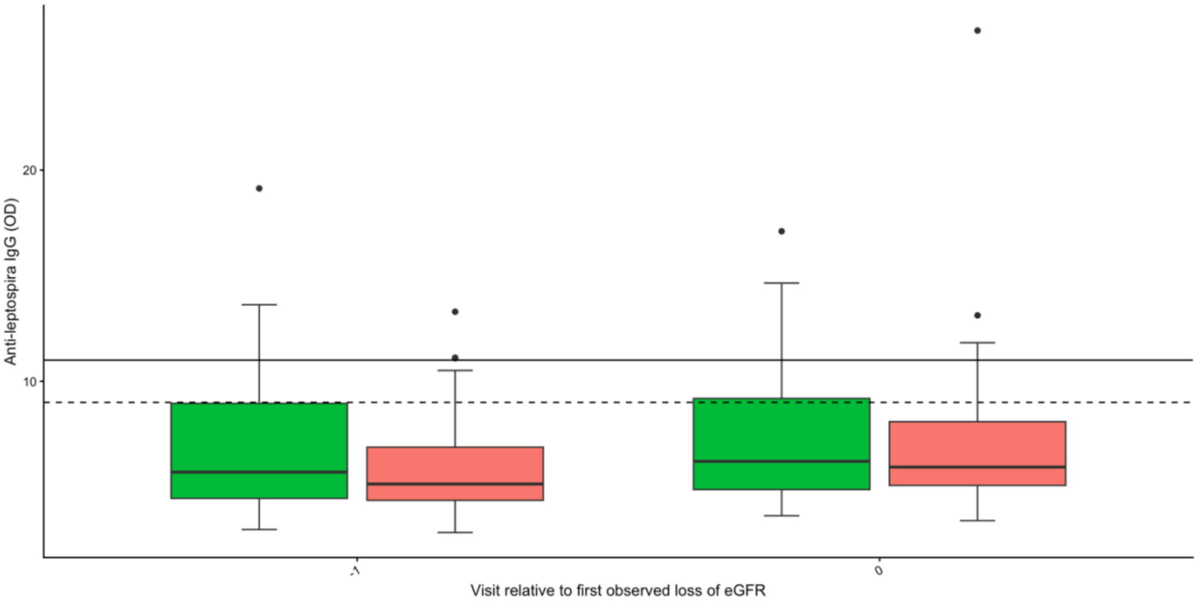

**Supplementary Figure 3 – Urinary CXCL9 levels in incident cases and controls**

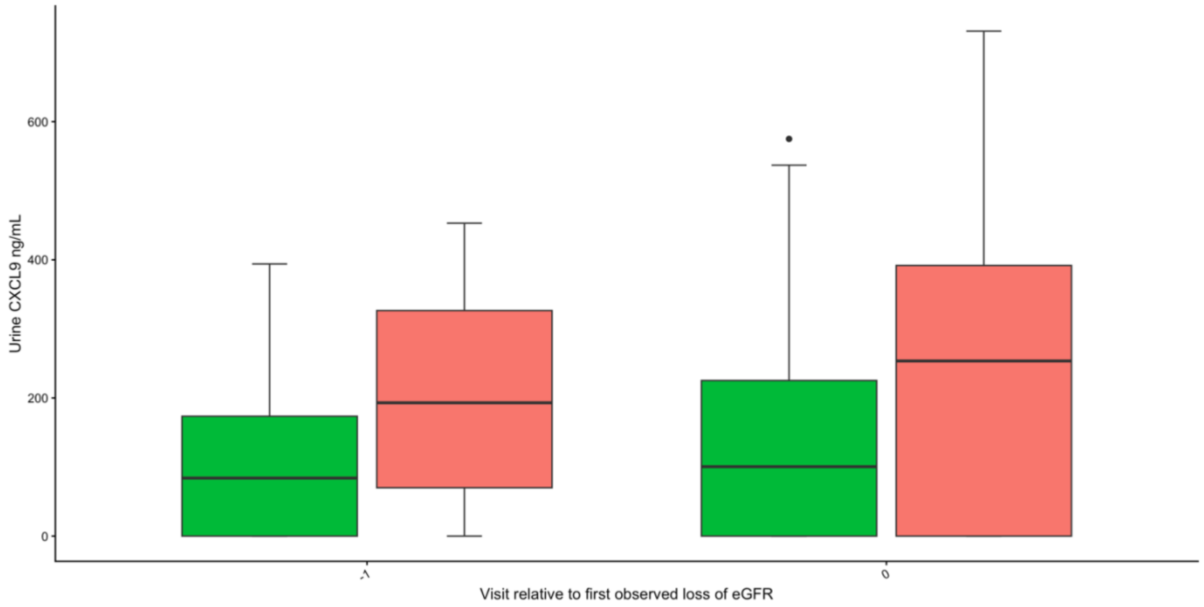

**Supplementary Figure 4 - Spaghetti plots of serum total IgE trajectories in the replication cohort**

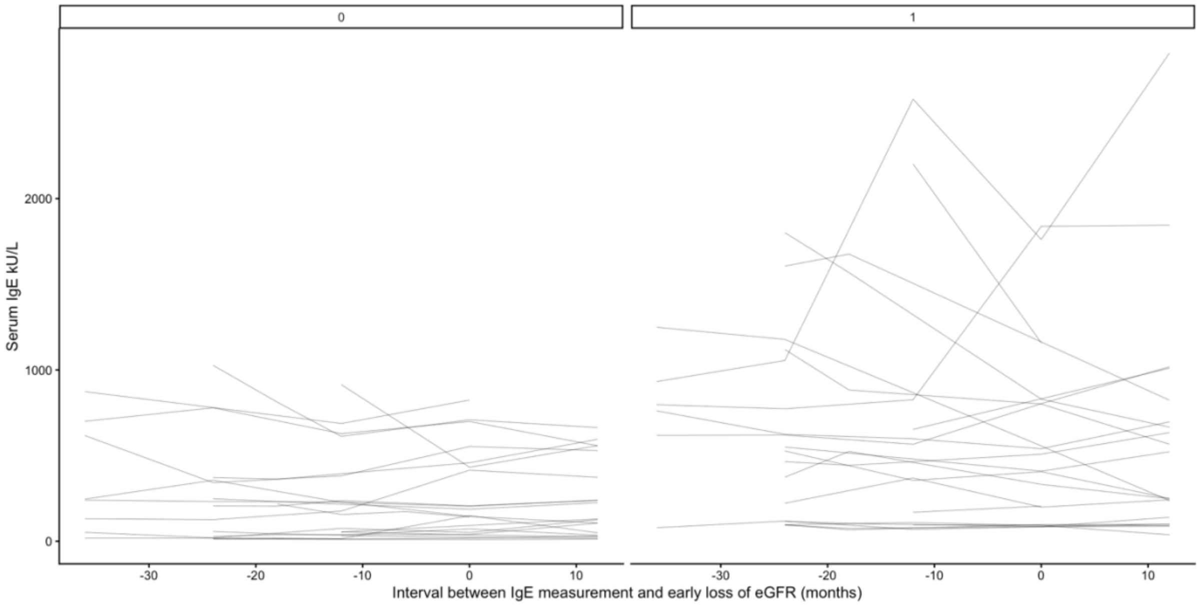

**Supplementary Figure 5 - Association between serum total IgE and eGFR by different time lags**

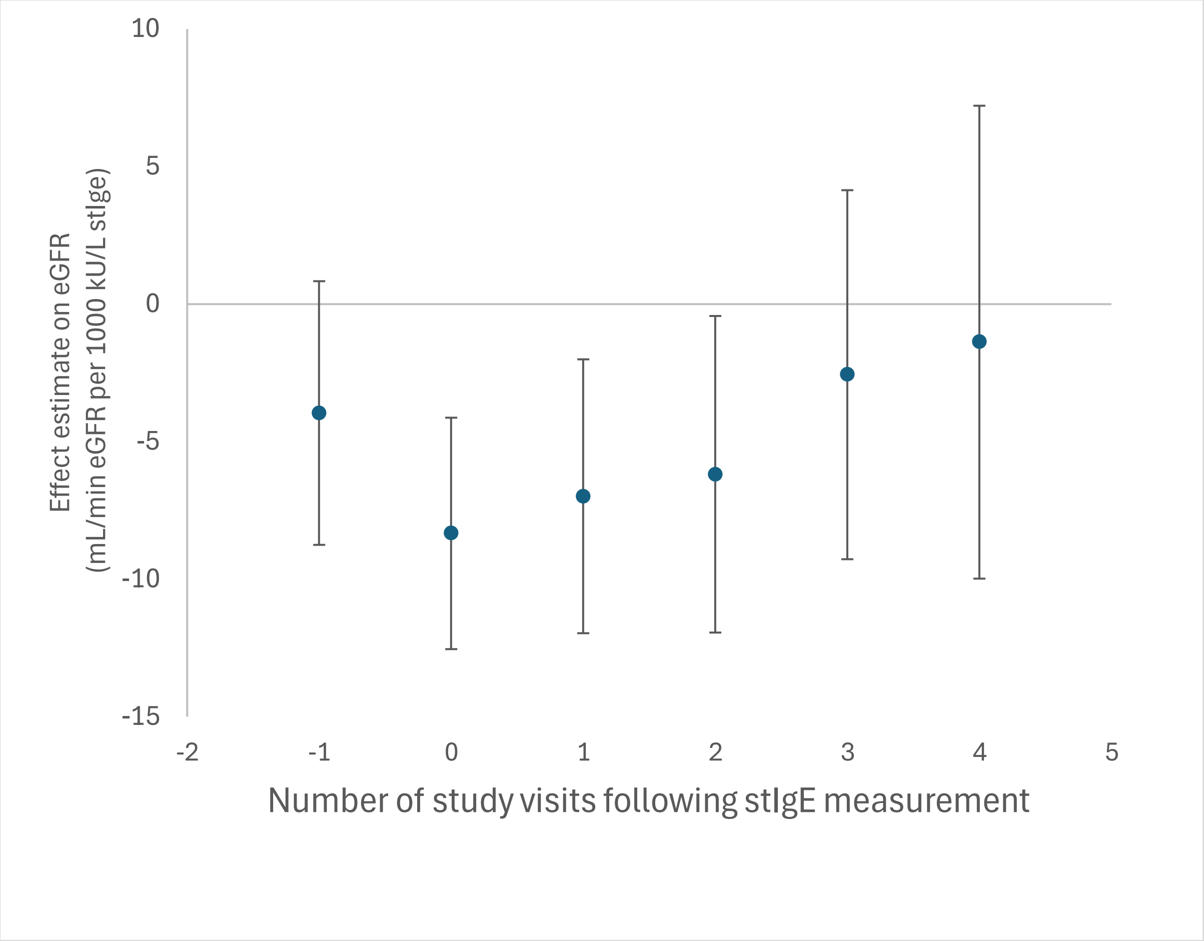

**STROBE Statement**

| | Item No. | Recommendation | Page No. | Relevant text from manuscript |
|---|---|---|---|---|
| **Title and abstract** | 1 | (*a*) Indicate the study's design with a commonly used term in the title or the abstract | 1 | |
| | | (*b*) Provide in the abstract an informative and balanced summary of what was done and what was found | 2 | |
| **Introduction** | | | | |
| Background/rationale | 2 | Explain the scientific background and rationale for the investigation being reported | 4 | |
| Objectives | 3 | State specific objectives, including any prespecified hypotheses | 4 | |
| **Methods** | | | | |
| Study design | 4 | Present key elements of study design early in the paper | 4 | |
| Setting | 5 | Describe the setting, locations, and relevant dates, including periods of recruitment, exposure, follow-up, and data collection | 4, S2 | |
| Participants | 6 | (*a*) *Cohort study*—Give the eligibility criteria, and the sources and methods of selection of participants. Describe methods of follow-up<br>*Case-control study*—Give the eligibility criteria, and the sources and methods of case ascertainment and control selection. Give the rationale for the choice of cases and controls<br>*Cross-sectional study*—Give the eligibility criteria, and the sources and methods of selection of participants | S2 | |
| | | (*b*) *Cohort study*—For matched studies, give matching criteria and number of exposed and unexposed<br>*Case-control study*—For matched studies, give matching criteria and the number of controls per case | S2 | |
| Variables | 7 | Clearly define all outcomes, exposures, predictors, potential confounders, and effect modifiers. Give diagnostic criteria, if applicable | S2 | |

| | | | |
|---|---|---|---|
| Data sources/ measurement | 8* | For each variable of interest, give sources of data and details of methods of assessment (measurement). Describe comparability of assessment methods if there is more than one group | S3-4 |
| Bias | 9 | Describe any efforts to address potential sources of bias | NA |
| Study size | 10 | Explain how the study size was arrived at | NA |

Continued on next page

| | | | |
|---|---|---|---|
| Quantitative variables | 11 | Explain how quantitative variables were handled in the analyses. If applicable, describe which groupings were chosen and why | 4, S3-4 |
| Statistical methods | 12 | (*a*) Describe all statistical methods, including those used to control for confounding | 4, S3-4 |
| | | (*b*) Describe any methods used to examine subgroups and interactions | |
| | | (*c*) Explain how missing data were addressed | |
| | | (*d*) *Cohort study*—If applicable, explain how loss to follow-up was addressed<br>*Case-control study*—If applicable, explain how matching of cases and controls was addressed<br>*Cross-sectional study*—If applicable, describe analytical methods taking account of sampling strategy | |
| | | (*e*) Describe any sensitivity analyses | |
| **Results** | | | |
| Participants | 13* | (a) Report numbers of individuals at each stage of study—eg numbers potentially eligible, examined for eligibility, confirmed eligible, included in the study, completing follow-up, and analysed | 5, S3-S8 |
| | | (b) Give reasons for non-participation at each stage | S2 |
| | | (c) Consider use of a flow diagram | NA |
| Descriptive data | 14* | (a) Give characteristics of study participants (eg demographic, clinical, social) and information on exposures and potential confounders | Table 1 |
| | | (b) Indicate number of participants with missing data for each variable of interest | Table 1 |
| | | (c) *Cohort study*—Summarise follow-up time (eg, average and total amount) | Table 1 |
| Outcome data | 15* | *Cohort study*—Report numbers of outcome events or summary measures over time | |
| | | *Case-control study*—Report numbers in each exposure category, or summary measures of exposure | 5, Table 1 |
| | | *Cross-sectional study*—Report numbers of outcome events or summary measures | |
| Main results | 16 | (*a*) Give unadjusted estimates and, if applicable, confounder-adjusted estimates and their precision (eg, 95% confidence interval). Make clear which confounders were adjusted for and why they were included | Table 2 |

| | | |
|---|---|---|
| | (*b*) Report category boundaries when continuous variables were categorized | 5 |
| | (c) If relevant, consider translating estimates of relative risk into absolute risk for a meaningful time period | NA |

Continued on next page

| | | | |
|---|---|---|---|
| Other analyses | 17 | Report other analyses done—eg analyses of subgroups and interactions, and sensitivity analyses | 6, S3-S13 |
| **Discussion** | | | |
| Key results | 18 | Summarise key results with reference to study objectives | 6 |
| Limitations | 19 | Discuss limitations of the study, taking into account sources of potential bias or imprecision. Discuss both direction and magnitude of any potential bias | 6 |
| Interpretation | 20 | Give a cautious overall interpretation of results considering objectives, limitations, multiplicity of analyses, results from similar studies, and other relevant evidence | 6-7 |
| Generalisability | 21 | Discuss the generalisability (external validity) of the study results | NA |
| **Other information** | | | |
| Funding | 22 | Give the source of funding and the role of the funders for the present study and, if applicable, for the original study on which the present article is based | 8 |

*Give information separately for cases and controls in case-control studies and, if applicable, for exposed and unexposed groups in cohort and cross-sectional studies.

**Note:** An Explanation and Elaboration article discusses each checklist item and gives methodological background and published examples of transparent reporting. The STROBE checklist is best used in conjunction with this article (freely available on the Web sites of PLoS Medicine at http://www.plosmedicine.org/, Annals of Internal Medicine at http://www.annals.org/, and Epidemiology at http://www.epidem.com/). Information on the STROBE Initiative is available at www.strobe-statement.org.
